# Evaluation of Tumor Response to Adjuvant Treatments using an Ex Vivo Culture of Breast Carcinoma Spheroids in a Microfluidic Device

**DOI:** 10.1101/2021.05.19.21257378

**Authors:** Hamidreza Aboulkheyr Es, Amir Reza Aref, Lobat Granpayeh, Marzieh Ebrahimi, Hossein Baharvand

## Abstract

**Purpose:** Breast cancer is the leading cause of cancer-related death among women worldwide. Conventional chemotherapy is considered a clinical state of the art treatment; however, resistance or recurrence occurs among a considerable portion of these patients. Besides understanding the genomic alterations pattern of tumor cells and their association with drug resistance or response, the development of a reliable tumor models that reflect the major cellular and molecular features of tumors may aid with screening of candidate drugs and identification of appropriate treatment regimens. Here, we developed a simple and low-cost tumor model of breast cancer to screen library of chemotherapy agents in a pre-clinical setting.

**Methods:** we generated and cultured ex-vivo 3D culture of patient-derived tumor spheroids from both pre-treated primary and metastatic tumors using a partial digestion approach in a microfluidic device. We assessed chemotherapy response of the seven patient-derived breast tumor spheroids and expanded evaluation of drug sensitivity through molecular analysis of a small panel of genes.

**Results:** We observed various chemotherapy responses across primary and metastasis tumor samples. Interestingly, we demonstrated response to paclitaxel and doxorubicin and resistance to cisplatin in 2/3 metastatic tumor samples while most of the primary tumor were responsive to chemotherapy. Additionally, the expression of *PIK3CA* and loss of *PTEN* were associated to treatment resistance.

**Conclusion:** Our study suggests potential application of microfluidic-based cell culture technology coupled with patient derived tumor spheroids in prediction of treatment response in a personalized manner.

## 1 Introduction

Breast cancer is the most prevalent cancer in women and remains a significant cause of cancer-associated death despite attempts to provide effective therapies. Patients diagnosed with triple-negative breast cancer (TNBC) do not benefit from endocrine or targeted therapies, and conventional chemotherapy is still considered the clinical state of the art for this subtype [1]. The development of high-throughput genomics technologies (e.g., microarrays and next-generation sequencing) has revolutionized our understanding of cancer genetics and its impact on treatment response and paved new ways for personalized cancer treatment [2]. As usual, when a patient’s tumor genomic pattern is used, a set of markers should be selected and combined with clinical information to build reliable information to predict the outcome of the patient as a candidate for chemotherapy [3,4]. Because of the molecular and cellular heterogeneity of the tumor microenvironment, which can significantly affect the response to therapy and clinical outcomes, the link between functional genomics and pathological data to predict patient outcome is a major challenge [5,6]. To address this challenge, reliable patient-derived in vitro tumor models that aid screening of candidate drugs may enhance the predictive power of the drug response, particularly in the case of chemotherapy [7]. In this regard, various patient-derived tumor models have been developed that range from patient-derived cell lines to mouse models generated from patient samples. Although these suggested models have opened new ways for drug testing, numerous drawbacks limit broad application of their use as drug-screening tools in pre-clinical settings [8,9]. Thus, a critical need exists for more sophisticated and low-cost pre-clinical cancer models that recapitulate human tumor biology and predict real-time response to chemotherapy. Mounting evidence has highlighted the application of microfluidic technology as a pre-clinical tumor model after successful ex vivo culture of various types of patient-derived tumors and screening library of anticancer drugs, including chemotherapy [10–15]. Tumor gene profiling of a panel of genes has aided prediction of chemotherapy response in breast cancer [4]. However, for those patients who have an intermediate score between response and resistance to chemotherapy, prediction of treatment response to standard of care therapy is challenging. We hypothesize that, besides the genomic pattern of the tumor, a three-dimensional (3D) ex vivo culture of patient-derived tumor samples with intact cellular heterogeneity [15,16] placed in a microfluidic device could be used to screen candidate chemotherapy agents. This system could be used to facilitate prediction of drug response in this population of patients.

As a proof-of-concept, we successfully cultured patient-derived tumor spheroids in a previously developed microfluidic platform and screened spheroids against a small library of chemotherapy drugs including doxorubicin, paclitaxel, and cisplatin. The behaviors of the tumor spheroids to these chemotherapy drugs were evaluated by both microscopic and molecular pattern analyses.

## 2 Materials and Methods

### 2-1 Patient-derived explant spheroid preparation and microfluidic culture

In this experimental study, breast cancer tissues were collected between January 2017 and January 2018 from seven females diagnosed with breast cancer (average range 37-56) and after study approval by Farmanieh Hospital and Sina Oncologic Hospital, according to local authorities. All seven patients signed a written informed consent form to authorize the use of their tissues for this study. All study procedures were conducted in accordance with the ethical standards approved by Royan Institute Ethical Committee (94000024) and the 1964 Helsinki Declaration and its later amendments or comparable ethical standards. Each patient’s histopathology information that included tumor size and depth of invasion; lymph-vascular and perineural invasion; grade and clinical tumor, node, and metastasis were recorded and pathologically staged using the tumor-node-metastasis (TNM) staging system.

Fresh tumor specimens that were received in media (DMEM) and on ice, were minced in a 10 cm dish (on ice) using sterile forceps and a scalpel. The minced tumor was resuspended in DMEM supplemented with 4.5 mM glucose, 100 mM sodium pyruvate, and 1:100 penicillin-streptomycin (all from Life Technologies, Corning Cellgro, Manassas, VA, USA), 100 U/mL collagenase type I (Life Technologies, Carlsbad, CA, USA), and 15 mM HEPES (Roche, Indianapolis, IN, USA). The samples were pelleted and resuspended in 10–20 mL of culture medium. Red blood cells were removed from visibly bloody samples using RBC Lysis Buffer (BioLegend, San Diego, CA, USA). The samples were pelleted and then resuspended in fresh DMEM + 10% FBS and strained through 100 µm and 70 µm mesh filters to generate 70–100 µm spheroid fractions. These generated fractions were primarily used for the ex vivo culture.

An aliquot of the fraction was pelleted, resuspended in collagen (prepared in 10x PBS, with the pH adjusted by using NaOH; pH 7.0–7.5), embedded in a collagen matrix, and loaded into the central channel of a 3D microfluidic culture device (Fig. 1) (AIM Biotech). The devices were kept in the incubator for 30 minutes to ECM polymerization. The culture medium with or without the chemotherapy drugs were loaded into two side channels of device. The devices were incubated for 72 hours.

**Figure 1.**
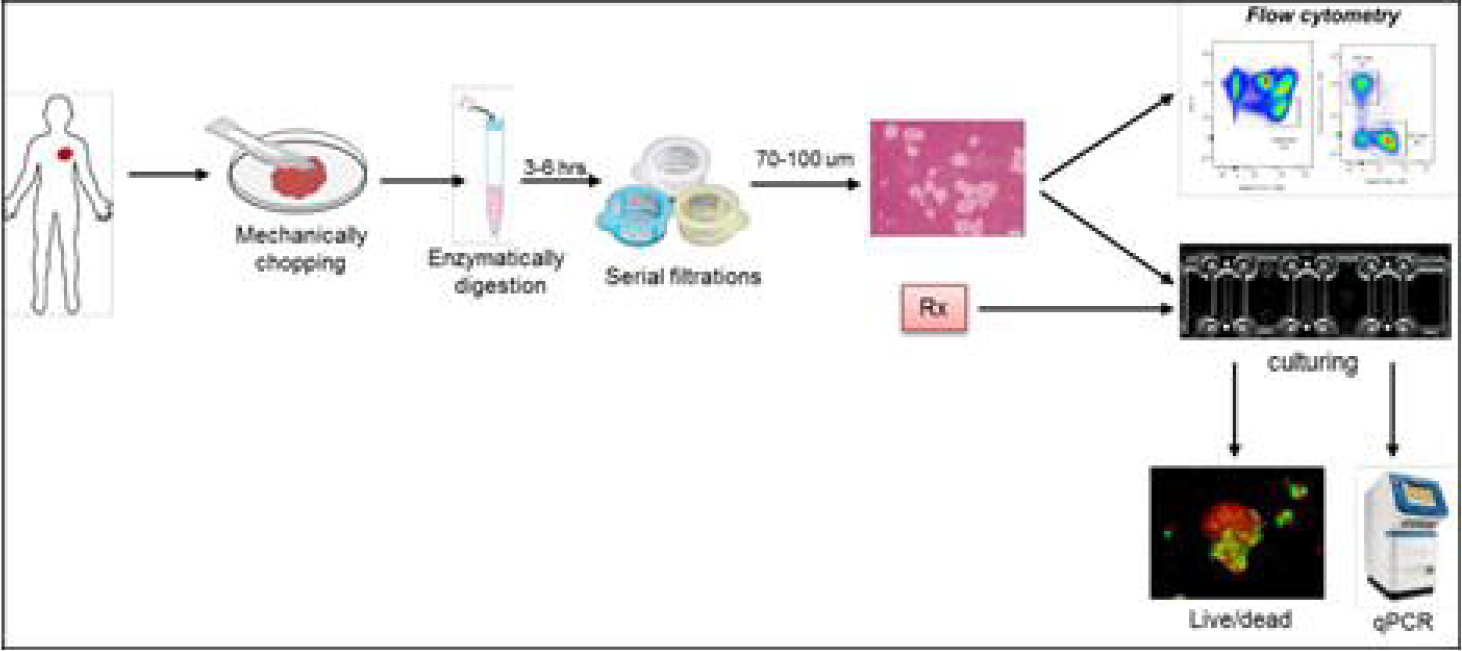
Schematic diagram of the experiment. The tumor specimens were received in transferring media immediately after surgery. These samples were washed twice with PBS, minced into small pieces, and partially digested with collagenase for four hours. Serial filtration was done to obtain tumor spheroids that ranged in size from 70–100 µm. One part of the isolated tumor spheroids was characterized using specific antibodies (EpCAM, CK14, CK 5/6, and CK 8/18). The other part was embedded in collagen type 1 and injected into the microfluidic device. Immediately after polymerization of the collagen, the device was filled with culture media that contained the chemotherapy medications and incubated for at least 72 hours.

### 2-2 Flow cytometry analysis

We assessed the expression levels of EpCAM, CK14, CK 5/6 (markers for basal-like cells), and CK 8/18 (marker for luminal-like cells) using a BD FACSCalibur flow cytometer. Briefly, the spheroids derived from fresh tumor tissues were enzymatically digested with 0.1% collagenase type I dissolved in DMEM base medium for 30 minutes at 37°C. The resulted single cells were centrifuged at 1000 RPM for 3 minutes and washed twise with PBS. The antibodies for EpCAM (Abcam, ab8666; FITC), CK14 (Abcam, ab192055; Alexa Fluor® 488), CK 5/6 (Abcam, ab193895; Alexa Fluor® 647), CK8 (Abcam, ab192468; Alexa Fluor® 647), and CK18 (Abcam, ab187573; Alexa Fluor® 488) was used according to the manufacturer’s protocol. The unstained cells were used as the negative control. The samples were analyzed with a DB-FACSCalibur and data analyzed with FlowJo software (version 10).

### 2-3 Live/dead immunostaining

Dual labeling with calcein-AM and propidium iodide (PI) was performed according to the manufacturer’s instructions. Briefly, the spheroids cultured in devices were imaged after labeling with calcein-AM (3 µm) and PI (4 µm) post-incubation for 10 minutes at 37°C. The images were captured by an inverted fluorescence microscope (Olympus IX71). Image deconvolution was performed using ImageJ software (version 1.8.0). Fluorescence quantitation (whole-device) was determined and used to quantify the relative number of live and dead cells by Olympus Cell-Sense Software.

### 2-4 Quantitative real-time PCR

The spheroids were extracted from the microfluidic device by enzymatic digestion of hydrogel with collagenase type I. We successfully isolated total RNA using an RnEasy Plus Microkit (Qiagen) according to the manufacturer’s instructions from all spheroids except BCP5. cDNA synthesis was performed with a Takara kit. Supplementary Table 1 lists the primers used in the present study.

### 2-5 Statistical analysis

All graphs and statistical analyses were performed using GraphPad/Prism (version 8.0). The t-test was performed for significant measurements. The P-values <0.05 was considered for statistical significance.

## 3 Results

### 3-1 Patient-derived spheroid preparation and characterization

In this study, we determined the possibility of culturing patient derived breast tumor spheroids from both neo-adjuvant and adjuvant treated tumors by adapting a previously designed 3D microfluidic multichannel device [17,18] (Fig. 1). The tumor specimens from seven patients with a mean age of 48 ± 8.04 years were used in this study. All tumors were classified as invasive ductal carcinoma (IDC). Immunohistochemical studies of estrogen receptor (ER) and progesterone receptor (PR) expressions were performed on the surgically resected tumor tissue samples based on standard methods. Tissue specimens from three patients were diagnosed as ER, PR and HER2 positive and four patients had undergone neoadjuvant therapy before sampling (Table 1).

**Table 1.**
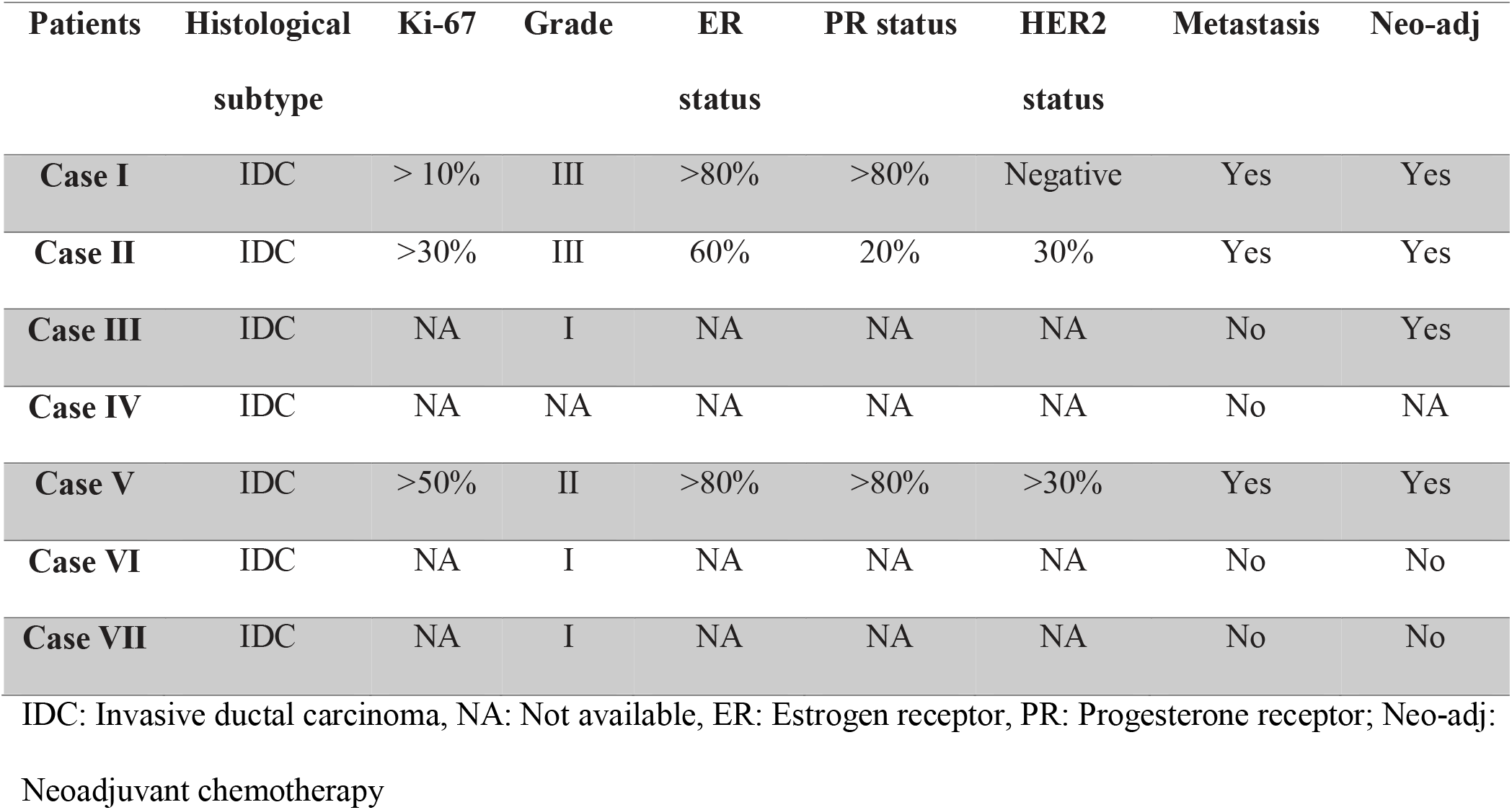
Clinicopathological features of the breast cancer patients.

| **Patients** | **Histological** | **Ki-67** | **Grade** | **ER** | **PR status** | **HER2** | **Metastasis** | **Neo-adj** |
| --- | --- | --- | --- | --- | --- | --- | --- | --- |
|  | **subtype** |  |  | **status** |  | **status** |  |  |
| **Case I** | IDC | > 10% | III | >80% | >80% | Negative | Yes | Yes |
| **Case II** | IDC | >30% | III | 60% | 20% | 30% | Yes | Yes |
| **Case III** | IDC | NA | I | NA | NA | NA | No | Yes |
| **Case IV** | IDC | NA | NA | NA | NA | NA | No | NA |
| **Case V** | IDC | >50% | II | >80% | >80% | >30% | Yes | Yes |
| **Case VI** | IDC | NA | I | NA | NA | NA | No | No |
| **Case VII** | IDC | NA | I | NA | NA | NA | No | No |

IDC: Invasive ductal carcinoma, NA: Not available, ER: Estrogen receptor, PR: Progesterone receptor; Neo-adj:
Neoadjuvant chemotherapy

To recapitulate the biology of the tumor microenvironment to model response of tumor to the chemotherapy drug, we need viable tumor tissue, an appropriate microfluidic device and a model of extracellular matrix to permit 3-dimentional culture. As number of tumor cells lose their viability within first hours of collection, it is essential to minimize duration of tumor specimen processing for culturing in device. The initial step in processing involves physical and enzymatic dissociation using a limited collagenase digestion. In this step we used a diluted concentration of the collagenase type I to avoid complete dissociation of the tumor specimens to single cells. Notably, the incubation time required to partially digestion of the mined tumor specimens with type I collagenase varies from sample to sample. Generally, those patient with a neo-adjuvant treatment regimen background represented a fibrous tumor compared to patients without an initial chemotherapy treatment before surgery.

Following physical and enzymatic dissociation, the minced and digested tumor specimen contains a mixture of macroscopic undigested tissue, spheroids, and single cells (Fig. 2). Spheroids are isolated following passage of the dissociated specimen over a series of filters (100μm and 70 μm). The enriched spheroids divided into three fractions, one fraction was selected for short term culturing in a ultra-low attached condition (Fig. 2), while the rest of two fraction used subsequently is used for molecular characterization and ex-vivo culture in microfluidic device.

**Figure 2:**
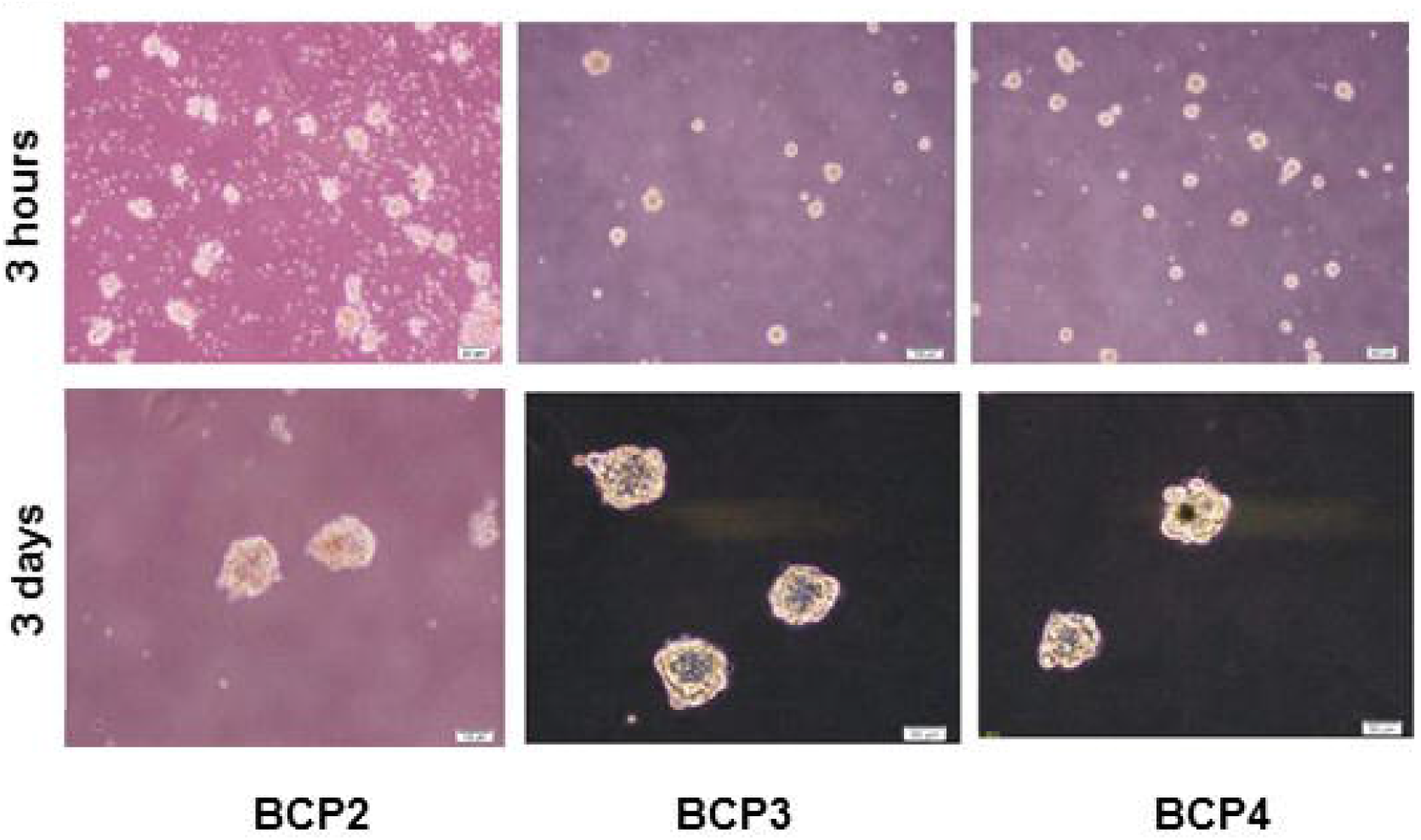
Generation of patient-derived spheroids. The tumor samples were enzymatically digested for three hours (upper panel; scale bar: 200 µm). Serial filtration was performed to capture spheroids that ranged from 70–100 µm. The isolated spheroids were cultured with complete medium in ultra-low attached conditions for three days (bottom panel; scale bar: 50 µm).

Morphologically, all isolated spheroids were round and compact. After culturing for three days in an ultra-low attached condition, we observed necrotic areas in the core of the spheroids (Fig. 2). Considering heterogeneity of the tumor cells, next we performed flow cytometry analysis for the expressions of EpCAM, CK14, and CK 5/6 (markers for basal-like cells), and CK 8/18 (marker for luminal-like cells) on fraction of patient derived spheroids with the diameter between 70-100 μm. The quantitative assessment of spheroids exhibited heterogeneity of expression of selected markers, particularly CK 8/18. All seven patient-derived spheroids expressed CK 8/18 and CK 5/14, while only four of seven spheroids expressed EpCAM (Fig. 3A, B). These data indicate presence of a heterogeneous population of cells even after course of treatment (Fig. 3B; BCP1, BCP2, BCP3, and BCP5).

**Figure 3:**
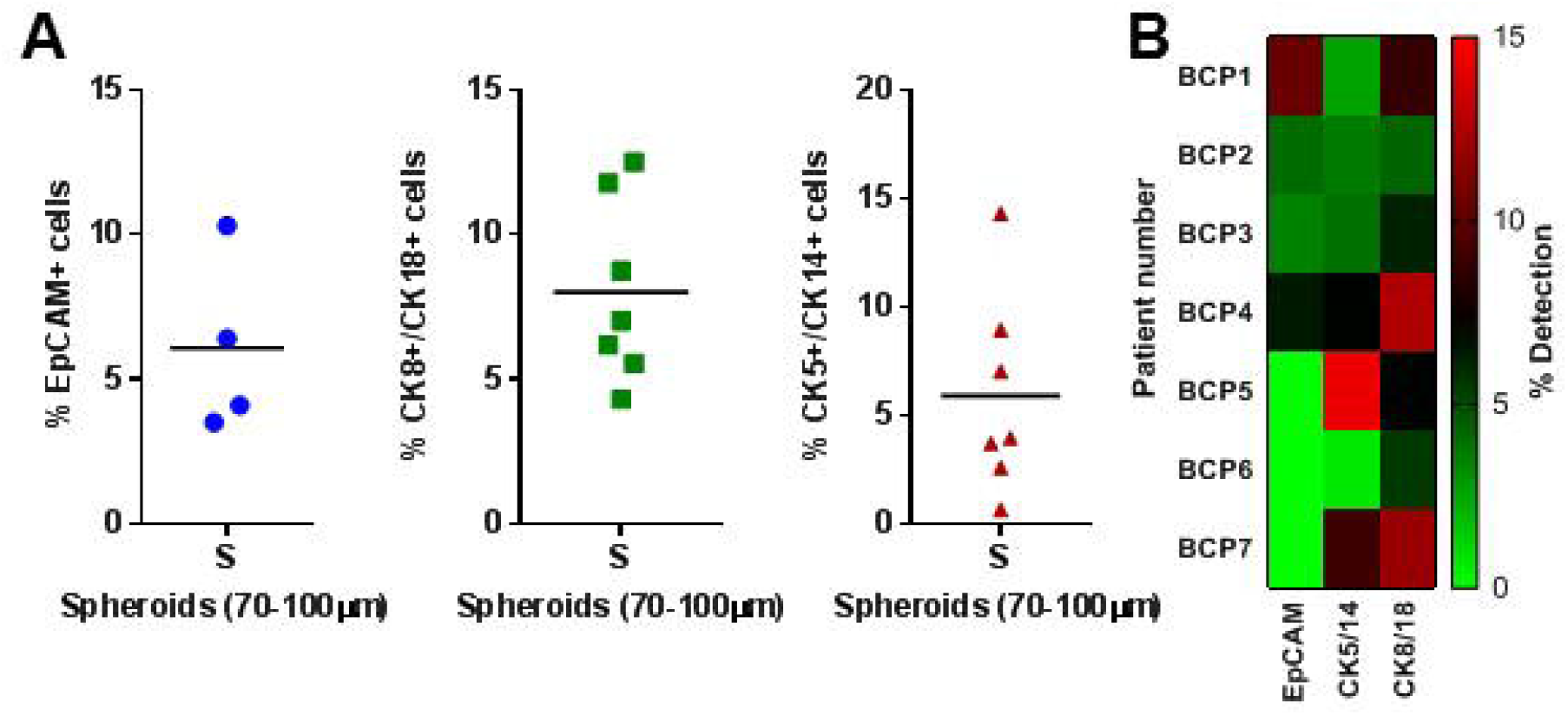
Flow cytometry analysis to quantify expression of breast cancer markers in patient-derived spheroids. The 70–100 µm spheroids were digested to achieve single cells for evaluation of human cytokeratin (CK) 5/14, CK 8/18 and EpCAM (A). Each dot represents an individual patient sample. (B) The heatmap represents an overview of CK 5/14, CK 8/18, and EpCAM expressions across seven samples.

### 3-2 Response of patient-derived spheroids to chemotherapy

Adjuvant chemotherapy as standard of care protocol for breast cancer can consist of paclitaxel and doxorubicin alone or in combination with cisplatin. As proof of concept, we screened patient-derived spheroids cultured in microfluidic devices against paclitaxel, doxorubicin, and cisplatin. We selected the medication for each sample in accordance with the physician’s prescription for each patient. Four samples were treated with doxorubicin (1 µm) and paclitaxel (0.1 µm) for 72 hours (Fig. 4). We measured the response of the spheroids to the selected drugs by live/dead staining of these spheroids in the devices. We found a significant response in three of four tumor-derived spheroids to paclitaxel (Fig. 4A-C, E). Spheroids derived from patient 5 were resistant to paclitaxel (Fig. 4D, E). In line with the microscopic observations, molecular analysis of the responsive spheroids (BCP2, BCP3, and BCP4) showed reduced expressions of *MKI67* and *PIK3CA*, and increased expressions of *P53* and *PTEN*. In some samples, *BCL2* had increased expression compared to the untreated group (Fig. 4A-C, E; left panel). Interestingly, the BCP5 samples had increased expressions of *TP53* and *BCL2* prior to therapy; however, there was reduced *PTEN* expression along with increased levels of *MKI67* and *PIK3CA* (Fig. 4D, 4E). In addition to these, different response to doxorubicin was observed in the patients’ samples. Spheroids derived from patients 2 and 4 (BCP2 and BCP4) showed more sensitivity to doxorubicin. A similar pattern of expression in the selected genes was observed in addition to the microscopic images (Fig. 4A, 4C, 4E). Interestingly, BCP3 spheroids presented an intermediate behavior of resistance or response to treatment in which an increase in expression of *TP53, PTEN*, and *MKI67*was recorded (Fig. 4B, 4E). Similar to BCP3, results from BCP5 suggested a mild resistance of the spheroids (Fig. 4D, 4E). In addition to the immunofluorescent data, reduced expression of *PTEN* and *MKI67* along with increased expression of *PIK3CA* indicated that these tumor cells were resistant to doxorubicin (Fig. 4D, 4E).

**Figure 4:**
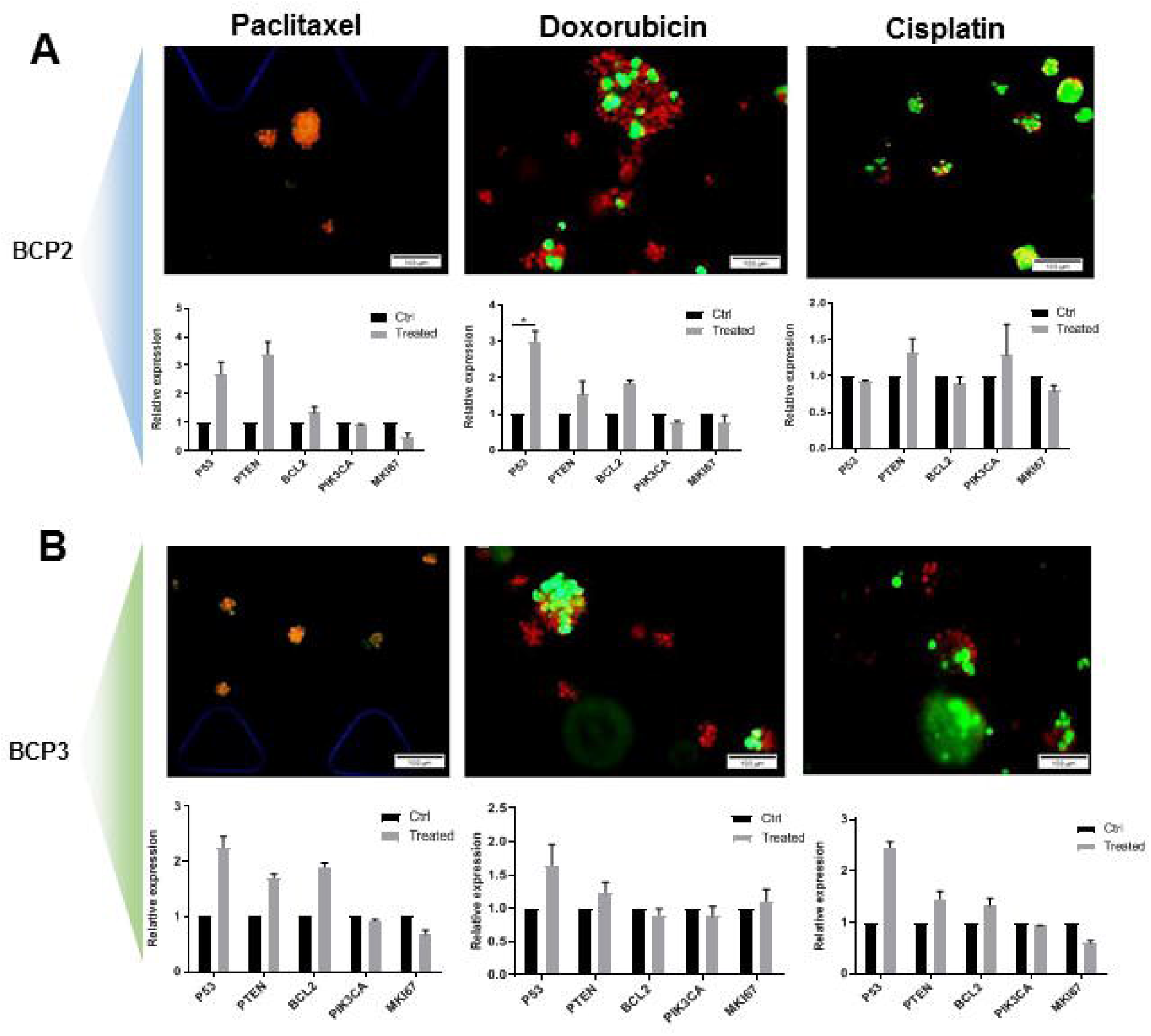
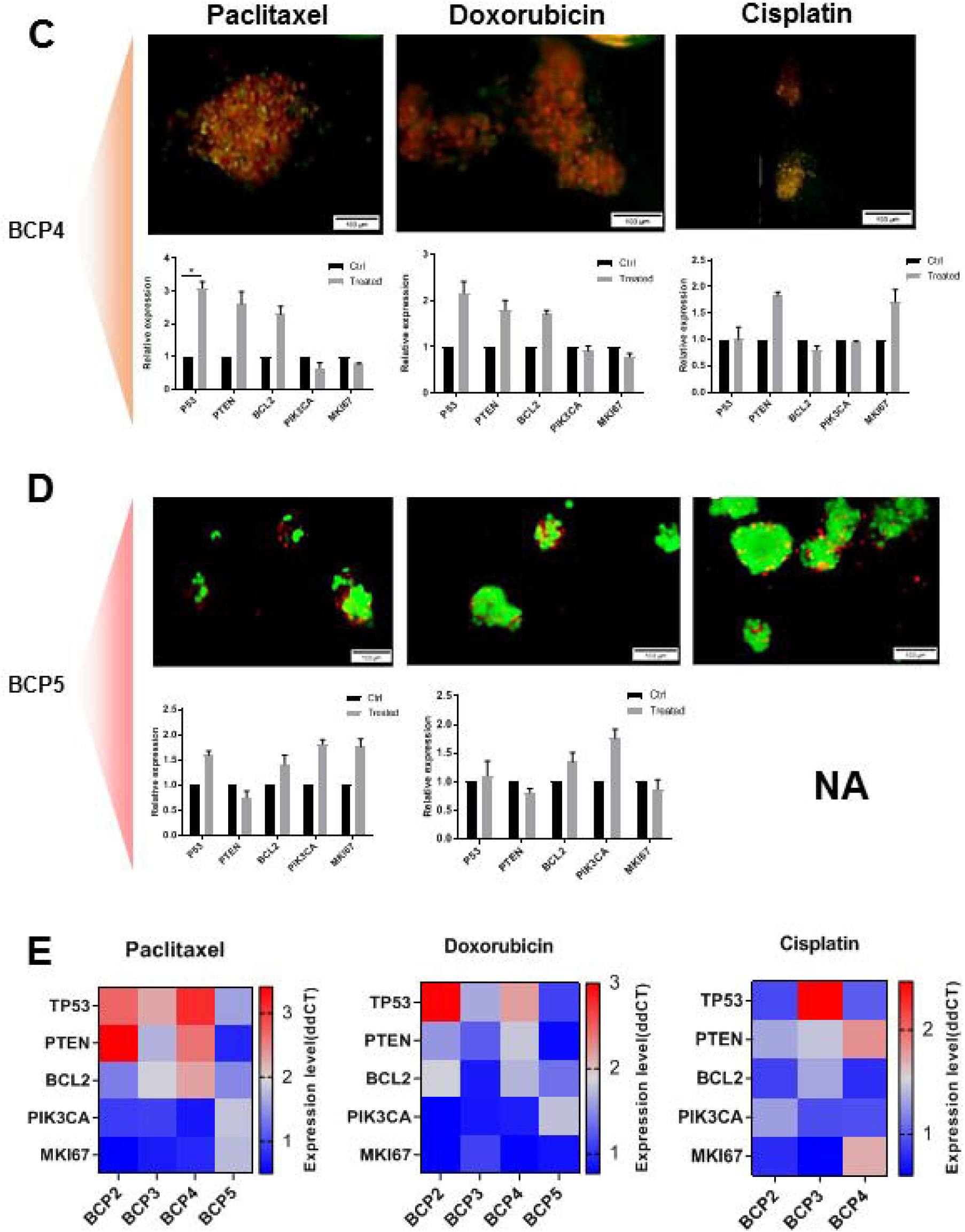
Sensitivity and resistance to the chemotherapy agents in the ex vivo tumor derived spheroids. The breast cancer spheroids derived from patients 2-5 were screened against paclitaxel, doxorubicin, and cisplatin. The live cells stained green with calcin-AM and dead cells stained red with propidium iodide (PI). Microscopic images show the sensitivity of the ex vivo spheroids to paclitaxel and doxorubicin in the BCP2, BCP3, and BCP4 samples 72 hours after treatment. A resistance-like behavior to cisplatin group was observed in all samples, except for BCP3. BCP5 was resistant to all of the screened drugs after 72 hours as indicated by the increased numbers of live cells (green area). Molecular typing was done for all treated spheroids and the results were compared with the untreated group. In support of the microscopic images, molecular analysis revealed that the expression levels of *P53, PTEN*, and *BCL2* increased in parallel with decreased proliferation of related genes as a result of apoptosis induction in the spheroids in tumor samples sensitive to paclitaxel (BCP2-4) and doxorubicin (BCP2 and 4). A partial response to doxorubicin was observed with expressions of *P53* and *MKI67* in BCP3. Spheroids derived from BCP5 showed resistance to paclitaxel and partial sensitivity to doxorubicin, which was accompanied by increased expression of *P53*, with increased cell proliferation. For cisplatin, all samples were resistant, except for BCP3 as determined by the expression levels of *P53* and *PIK3CA*. (scale bar: 100 µm). The heatmap illustrates a combined view of the expressions of the selected gene panel in different samples from each treatment group. NA: not available.

We screened all spheroids against cisplatin (Fig. 4A-E). The BCP2, BCP4, and BCP5 spheroids were resistant to cisplatin as shown by the results of the both live/dead assays and molecular analysis (Fig. 4A, 4C-E). The samples derived from patient 3 (BCP3) were sensitive to cisplatin. The molecular measurement was in line with the microscopic observation of elevated expression of *TP53* and a reduced expression of the proliferation-related gene, *MKI67* (Fig. 4B, 4E). Taken together, the sensitive spheroids to paclitaxel and doxorubicin had significantly increased levels of *P53* and *PTEN*, and, in some cases, *BCL2* (Fig. 4), whereas the increase in *PIK3CA* level alone or in conjunction with *MKI67* and reduction of *PTEN* in some spheroids was associated with resistance to paclitaxel, doxorubicin, and cisplatin (Fig. 4).

## 4 Discussion

Prediction of drug response in cancer patients is a major challenge in the clinic. Various tumor models have been proposed to overcome this challenge [6]. Patient-derived tumor xenografts models (PDX) are extensively used for drug response prediction, but variations in successful engraftment are limited to the engraftment from advanced cancer stages and the extended period of time between cancer diagnosis and creation of the xenografts. This limits the application of PDX models as preclinical tumor models [9,19–23]. Although the advent of mouse PDX models has opened up the possibility of tailoring drugs to a tumor with a specific genetic lesion; however, in practice, these models are expensive and human tumor stromal cells can be replaced by mice stromal cells resulting different drug response[9,24].

Recently, various preclinical studies have cultured patient-derived tumor organoids in microfluidic devices for various types of cancers, particularly lung adenocarcinoma and melanoma [14,15,20,25]. Here, we adapted and developed a rapid, inexpensive platform of an in vitro tumor model that can be used in personalized medicine. This tumor model relies on an ex vivo 3D culture of patient-derived breast tumor tissues in microfluidic devices and has the capability to predict drug response in a clinical and pre-clinical setting.

In line with this, a large body of evidence suggested the application of the microfluidic system in drug screening and personalized cancer medicine[10,12]. Considering the role of drug screening platforms for measuring and prediction of synergy and/or additive effects of two drugs, microfluidic culture systems can show the synergistic effects of two drugs, while in conventional methods, additive effects are observed [26]. In these devices, an uniform distribution of materials including drug or media is observed because of the architecture and nature of the microfluidic devices and the culture of a collagen embedded tumor spheroid in this device [16,27,28]. We observed a mild effect of cultivation on generated spheroids after 24 hours of culture. Our preference was to examine drug responses immediately after generating spheroids to minimize any effects of the culture.

We selected a small panel of recurrence genes (*P53, PTEN, BCL2, PIK3CA*, and *MKI67*) based on previously published high-throughput cancer genome analysis in breast cancer and analyzed patient-derived organoids after treatment at the molecular level [28–33]. In agreement with previous works, we found that increases in the levels of *P53, PTEN*, and *BCL2* in some samples could lead to a significant response to paclitaxel, doxorubicin, and cisplatin in breast cancer [33– 35]. The *P53* is one of the well-studied tumor suppressor genes. Its mutations caus tumor development, but its expression positively correlates with good prognosis in breast cancer patients [36,37].

A long with the *P53*, the role of *PTEN* in drug response and resistance in breast cancer has been extensively reported [36,38], in which elevated expression of *PTEN* is associated with better chemotherapy response resulting in sensitive behavior. However, loss of *PTEN* and expression of *PIK3CA* in breast cancer is a marker of insensitivity to standard therapy [39]. In parallel with other studies, we found that over-expression of *MKI67* alone or combined with *PIK3CA* could determine drug responsiveness in several cancers, including breast cancer [38,40]. Although the molecular type was in line with live and dead staining in the present study, variations in the molecular results made it difficult to predict patient outcome. Indeed, with a simple test on tumor-derived spheroids in a microfluidic device, we could easily predict the patient response to chemotherapy in a short period of time (within 3 days) without an analysis of different patterns of genes.

In summary, the microfluidic ex vivo culture platform provides a simple and easy patient-relevant model system for preclinical evaluation of novel anticancer agents. The platform can be combined with an analysis of the expression patterns tumor specimens to provide a reliable platform for personalized drug treatment in breast cancer patients. This platform is inexpensive, rapid, and achievable within an integrated cancer translational research setting.

## Data Availability

Not available

## Acknowledgements

This study was founded by grants awarded by Royan Institute, and the Iran National Science Foundation (INSF), [Grant No. 96001316] to H.B.

---

**Supplementary Table 1.**
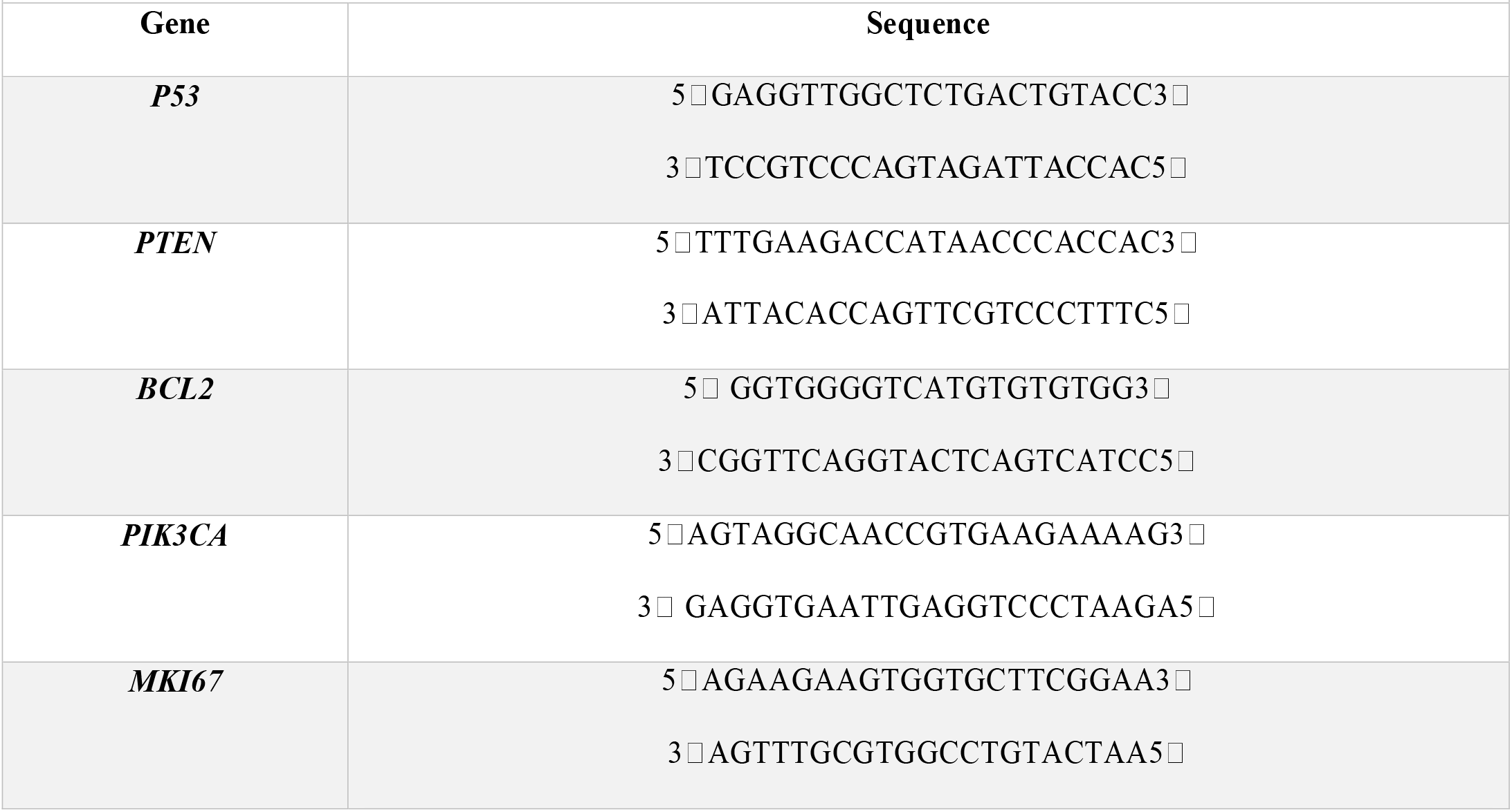
Primer sequences used in the present study.

## References

1. C. Denkert, C. Liedtke, A. Tutt, and G. von Minckwitz, Lancet 389, 2430 (2017).

2. A. a. Friedman, A. Letai, D. E. Fisher, and K. T. Flaherty, Nat. Rev. Cancer 15, 747 (2015).

3. C. Kim, R. Gao, E. Sei, R. Brandt, J. Hartman, T. Hatschek, N. Crosetto, T. Foukakis, and N. E. Navin, Cell 173, 879 (2018).

4. Y. Z. Jiang, D. Ma, C. Suo, J. Shi, M. Xue, X. Hu, Y. Xiao, K. Da Yu, Y. R. Liu, Y. Yu, Y. Zheng, X. Li, C. Zhang, P. Hu, J. Zhang, Q. Hua, J. Zhang, W. Hou, L. Ren, D. Bao, B. Li, J. Yang, L. Yao, W. J. Zuo, S. Zhao, Y. Gong, Y. X. Ren, Y. X. Zhao, Y. S. Yang, Z. Niu, Z. G. Cao, D. G. Stover, C. Verschraegen, V. Kaklamani, A. Daemen, J. R. Benson, K. Takabe, F. Bai, D. Q. Li, P. Wang, L. Shi, W. Huang, and Z. M. Shao, ancer Cell (2019).

5. D. Tuveson and H. Clevers, Science (80-.). 364, 952 (2019).

6. H. Aboulkheyr Es, L. Montazeri, A. R. Aref, M. Vosough, and H. Baharvand, Trends Biotechnol. 36, 358 (2018).

7. F. Weeber, S. N. Ooft, K. K. Dijkstra, and E. E. Voest, Cell Chem. Biol. (2017).

8. H. F. Tsai, A. Trubelja, A. Q. Shen, and G. Bao, J. R. Soc. Interface (2017).

9. M. Hidalgo, F. Amant, A. V. Biankin, E. Budinsk??, A. T. Byrne, C. Caldas, R. B. Clarke, S. de Jong, J. Jonkers, G. M. M??landsmo, S. Roman-Roman, J. Seoane, L. Trusolino, and A. Villanueva, Cancer Discov. 4, 998 (2014).

10. A. Sontheimer-Phelps, B. A. Hassell, and D. E. Ingber, Nat. Rev. Cancer (2019).

11. K. P. Valente, S. Khetani, A. R. Kolahchi, A. Sanati-Nezhad, A. Suleman, and M. Akbari, Drug Discov. Today (2017).

12. F. Eduati, R. Utharala, D. Madhavan, U. P. Neumann, T. Longerich, T. Cramer, J. Saez-Rodriguez, and C. A. Merten, Nat. Commun. (2018).

13. M. Shang, R. H. Soon, C. T. Lim, B. L. Khoo, and J. Han, Lab Chip (2019).

14. M. Sade-Feldman, K. Yizhak, S. L. Bjorgaard, J. P. Ray, C. G. de Boer, R. W. Jenkins, D. J. Lieb, J. H. Chen, D. T. Frederick, M. Barzily-Rokni, S. S. Freeman, A. Reuben, P. J. Hoover, A.-C. Villani, E. Ivanova, A. Portell, P. H. Lizotte, A. R. Aref, J.-P. Eliane, M. R. Hammond, H. Vitzthum, S. M. Blackmon, B. Li, V. Gopalakrishnan, S. M. Reddy, Z. Cooper, C. P. Paweletz, D. A. Barbie, A. Stemmer-Rachamimov, K. T. Flaherty, J. A. Wargo, G. M. Boland, R. J. Sullivan, G. Getz, and N. Hacohen, Cell 175, 998 (2018).

15. A. R. Aref, M. Campisi, E. Ivanova, A. Portell, D. Larios, B. P. Piel, N. Mathur, C. Zhou, R. V. Coakley, A. Bartels, M. Bowden, Z. Herbert, S. Hill, S. Gilhooley, J. Carter, I. Cañadas, T. C. Thai, S. Kitajima, V. Chiono, C. P. Paweletz, D. A. Barbie, R. D. Kamm, and R. W. Jenkins, Lab Chip (2018).

16. A. R. Aref, R. Y. J. Huang, W. Yu, K. N. Chua, W. Sun, T. Y. Tu, J. Bai, W. J. Sim, I. K. Zervantonakis, J. P. Thiery, and R. D. Kamm, Integr. Biol. (United Kingdom) (2013).

17. A. Aref, E. Ivanova, E. Chambers, A. Portell, R. Jenkins, M. Lin, M. Xu, J. Haworth, M. Bahcall, and C. Paweletz, (2018).

18. A. R. Aref, M. Campisi, E. Ivanova, A. Portell, D. Larios, B. P. Piel, N. Mathur, C. Zhou, R. V. Coakley, A. Bartels, M. Bowden, Z. Herbert, S. Hill, S. Gilhooley, J. Carter Cañadas, T. C. Thai, S. Kitajima, V. Chiono, C. P. Paweletz, D. A. Barbie, R. D. Kamm, and R. W. Jenkins, Lab Chip 3129 (2018).

19. I. Holen, V. Speirs, B. Morrissey, and K. Blyth, Dis. Model. Mech. (2017).

20. F. Mattei, G. Schiavoni, A. De Ninno, V. Lucarini, P. Sestili, A. Sistigu, A. Fragale, M. Sanchez, M. Spada, A. Gerardino, F. Belardelli, L. Businaro, and L. Gabriele, J. Immunotoxicol. 11, 337 (2014).

21. P. Malaney, S. V. Nicosia, and V. Dav??, Cancer Lett. 344, 1 (2014).

22. M. Hidalgo, E. Bruckheimer, N. V Rajeshkumar, I. Garrido-Laguna, E. De Oliveira, B. Rubio-Viqueira, S. Strawn, M. J. Wick, J. Martell, and D. Sidransky, Mol. Cancer Ther. 10, 1311 (2011).

23. C. Pauli, B. D. Hopkins, D. Prandi, R. Shaw, T. Fedrizzi, A. Sboner, V. Sailer, M. Augello, L. Puca, R. Rosati, T. J. McNary, Y. Churakova, C. Cheung, J. Triscott, D. Pisapia, R. Rao, J. M. Mosquera, B. Robinson, B. M. Faltas, B. E. Emerling, V. K. Gadi, B. Bernard, O. Elemento, H. Beltran, F. Demichelis, C. J. Kemp, C. Grandori, L. C. Cantley, and M. A. Rubin, Cancer Discov. (2017).

24. R. W. Jenkins, A. R. Aref, P. H. Lizotte, E. Ivanova, S. Stinson, C. W. Zhou, M. Bowden, J. Deng, H. Liu, D. Miao, M. X. He, W. Walker, G. Zhang, T. Tian, C. Cheng, Z. Wei, S. Palakurthi, M. Bittinger, H. Vitzthum, J. W. Kim, A. Merlino, M. Quinn, C. Venkataramani, J. A. Kaplan, A. Portell, P. C. Gokhale, B. Phillips, A. Smart, A. Rotem, R. E. Jones, L. Keogh, M. Anguiano, L. Stapleton, Z. Jia, M. Barzily-Rokni, I. Cañadas, T. C. Thai, M. R. Hammond, R. Vlahos, E. S. Wang, H. Zhang, S. Li, G. J. Hanna, W. Huang, M. P. Hoang, A. Piris, J. P. Eliane, A. O. Stemmer-Rachamimov, L. Cameron, M. J. Su, P. Shah, B. Izar, M. Thakuria, N. R. LeBoeuf, G. Rabinowits, V. Gunda, S. Parangi, J. M. Cleary, B. C. Miller, S. Kitajima, R. Thummalapalli, B. Miao, T. U. Barbie, V. Sivathanu, J. Wong, W. G. Richards, R. Bueno, C. H. Yoon, J. Miret, M. Herlyn, L. A. Garraway, E. M. Van Allen, G. J. Freeman, P. T. Kirschmeier, J. H. Lorch, P. A. Ott, F. Stephen Hodi, K. T. Flaherty, R. D. Kamm, G. M. Boland, K. K. Wong, D. Dornan, C. P. Paweletz, and D. A. Barbie, Cancer Discov. (2018).

25. J. H. Tsui, W. Lee, S. H. Pun, J. Kim, and D. H. Kim, Adv. Drug Deliv. Rev. (2013).

26. Y. Shin, S. Han, J. S. Jeon, K. Yamamoto, I. K. Zervantonakis, R. Sudo, R. D. Kamm, and S. Chung, Nat. Protoc. (2012).

27. A. A., I. E., C. E., P. A., J. R., L. M., X. M., H. J., B. M., P. C., and J. P., Cancer Res. (2018).

28. N. Sachs, J. de Ligt, O. Kopper, E. Gogola, G. Bounova, F. Weeber, A. V. Balgobind, K. Wind, A. Gracanin, H. Begthel, J. Korving, R. van Boxtel, A. A. Duarte, D. Lelieveld, A. van Hoeck, R. F. Ernst, F. Blokzijl, I. J. Nijman, M. Hoogstraat, M. van de Ven, D. A. Egan, V. Zinzalla, J. Moll, S. F. Boj, E. E. Voest, L. Wessels, P. J. van Diest, S. Rottenberg, R. G. J. Vries, E. Cuppen, and H. Clevers, Cell (2018).

29. P. Razavi, M. T. Chang, G. Xu, C. Bandlamudi, D. S. Ross, N. Vasan, Y. Cai, C. M. Bielski, M. T. A. Donoghue, P. Jonsson, A. Penson, R. Shen, F. Pareja, R. Kundra, S. Middha, M. L. Cheng, A. Zehir, C. Kandoth, R. Patel, K. Huberman, L. M. Smyth, K. Jhaveri, S. Modi, T. A. Traina, C. Dang, W. Zhang, B. Weigelt, B. T. Li, M. Ladanyi, D. M. Hyman, N. Schultz, M. E. Robson, C. Hudis, E. Brogi, A. Viale, L. Norton, M. N. Dickler, M. F. Berger, C. A. Iacobuzio-Donahue, S. Chandarlapaty, M. Scaltriti, J. S. Reis-Filho, D. B. Solit, B. S. Taylor, and J. Baselga, Cancer Cell 34, 427 (2018).

30. A. C. Berger, A. Korkut, R. S. Kanchi, A. M. Hegde, W. Lenoir, W. Liu, Y. Liu, H. Fan, H. Shen, V. Ravikumar, A. Rao, A. Schultz, X. Li, P. Sumazin, C. Williams, P. Mestdagh, P. H. Gunaratne, C. Yau, R. Bowlby, A. G. Robertson, D. G. Tiezzi, C. Wang, D. Cherniack, A. K. Godwin, N. M. Kuderer, J. S. Rader, R. E. Zuna, A. K. Sood, A. J. Lazar, A. I. Ojesina, C. Adebamowo, S. N. Adebamowo, K. A. Baggerly, T. W. Chen, H. S. Chiu, S. Lefever, L. Liu, K. MacKenzie, S. Orsulic, J. Roszik, C. S. Shelley, Q. Song, C. P. Vellano, N. Wentzensen, S. J. Caesar-Johnson, J. A. Demchok, I. Felau, M. Kasapi, M. L. Ferguson, C. M. Hutter, H. J. Sofia, R. Tarnuzzer, Z. Wang, L. Yang, J. C. Zenklusen, J. (Julia) Zhang, S. Chudamani, J. Liu, L. Lolla, R. Naresh, T. Pihl, Q. Sun, Y. Wan, Y. Wu, J. Cho, T. DeFreitas, S. Frazer, N. Gehlenborg, G. Getz, D. I. Heiman, J. Kim, M. S. Lawrence, P. Lin, S. Meier, M. S. Noble, G. Saksena, D. Voet, H. Zhang, B. Bernard, N. Chambwe, V. Dhankani, T. Knijnenburg, R. Kramer, K. Leinonen, M. Miller, S. Reynolds, I. Shmulevich, V. Thorsson, W. Zhang, R. Akbani, B. M. Broom, Z. Ju, J. Li, H. Liang, S. Ling, Y. Lu, G. B. Mills, K. S. Ng, M. Ryan, J. Wang, J. N. Weinstein, J. Zhang, A. Abeshouse, J. Armenia, D. Chakravarty, W. K. Chatila, I. de Bruijn, J. Gao, B. E. Gross, Z. J. Heins, R. Kundra, K. La, M. Ladanyi, A. Luna, M. G. Nissan, A. Ochoa, S. M. Phillips, E. Reznik, F. Sanchez-Vega, C. Sander, N. Schultz, R. Sheridan, S. O. Sumer, Y. Sun, B. S. Taylor, P. Anur, M. Peto, P. Spellman, C. Benz, J. M. Stuart, C. K. Wong, D. N. Hayes, J. S. Parker, M. D. Wilkerson, A. Ally, M. Balasundaram, D. Brooks, R. Carlsen, E. Chuah, N. Dhalla, R. Holt, S. J. M. Jones, K. Kasaian, D. Lee, Y. Ma, M. A. Marra, M. Mayo, R. A. Moore, A. J. Mungall, K. Mungall, S. Sadeghi, J. E. Schein, P. Sipahimalani, A. Tam, N. Thiessen, K. Tse, T. Wong, R. Beroukhim, C. Cibulskis, S. B. Gabriel, G. F. Gao, G. Ha, M. Meyerson, S. E. Schumacher, J. Shih, M. H. Kucherlapati, R. S. Kucherlapati, S. Baylin, L. Cope, L. Danilova, M. S. Bootwalla, P. H. Lai, D. T. Maglinte, D. J. Van Den Berg, D. J. Weisenberger, J. T. Auman, S. Balu, T. Bodenheimer, C. Fan, K. A. Hoadley, A. P. Hoyle, S. R. Jefferys, C. D. Jones, S. Meng, P. A. Mieczkowski, L. E. Mose, A. H. Perou, C. M. Perou, J. Roach, Y. Shi, J. V. Simons, T. Skelly, M. G. Soloway, D. Tan, U. Veluvolu, T. Hinoue, P. W. Laird, W. Zhou, M. Bellair, K. Chang, K. Covington, C. J. Creighton, H. Dinh, H. V. Doddapaneni, L. A. Donehower, J. Drummond, R. A. Gibbs, R. Glenn, W. Hale, Y. Han, J. Hu, V. Korchina, S. Lee, L. Lewis, W. Li, X. Liu, M. Morgan, D. Morton, D. Muzny, J. Santibanez, M. Sheth, E. Shinbrot, L. Wang, M. Wang, D. A. Wheeler, L. Xi, F. Zhao, J. Hess, E. L. Appelbaum, M. Bailey, M. G. Cordes, L. Ding, C. C. Fronick, L. A. Fulton, R. S. Fulton, C. Kandoth, E. R. Mardis, M. D. McLellan, C. A. Miller, H. K. Schmidt, R. K. Wilson, D. Crain, E. Curley, J. Gardner, K. Lau, D. Mallery, S. Morris, J. Paulauskis, R. Penny, C. Shelton, T. Shelton, M. Sherman, E. Thompson, P. Yena, J. Bowen, J. M. Gastier-Foster, M. Gerken, K. M. Leraas, T. M. Lichtenberg, N. C. Ramirez, L. Wise, E. Zmuda, N. Corcoran, T. Costello, C. Hovens, A. L. Carvalho, A. C. de Carvalho, J. H. Fregnani, A. Longatto-Filho, R. M. Reis, C. Scapulatempo-Neto, H. C. S. Silveira, D. O. Vidal, A. Burnette, J. Eschbacher, B. Hermes, A. Noss, R. Singh, M. L. Anderson, P. D. Castro, M. Ittmann, D. Huntsman, B. Kohl, X. Le, R. Thorp, C. Andry, E. R. Duffy, V. Lyadov, O. Paklina, G. Setdikova, A. Shabunin, M. Tavobilov, C. McPherson, R. Warnick, R. Berkowitz, D. Cramer, C. Feltmate, N. Horowitz, A. Kibel, M. Muto, C. P. Raut, A. Malykh, J. S. Barnholtz-Sloan, W. Barrett, K. Devine, J. Fulop, Q. T. Ostrom, K. Shimmel, Y. Wolinsky, A. E. Sloan, A. De Rose, F. Giuliante, M. Goodman, B. Y. Karlan, C. H. Hagedorn, J. Eckman, J. Harr, J. Myers, K. Tucker, L. A. Zach, B. Deyarmin, H. Hu, L. Kvecher, C. Larson, R. J. Mural, S. Somiari, A. Vicha, T. Zelinka, J. Bennett, M. Iacocca, B. Rabeno, P. Swanson, M. Latour, L. Lacombe, B. Têtu, A. Bergeron, M. McGraw, S. M. Staugaitis, J. Chabot, H. Hibshoosh, A. Sepulveda, T. Su, T. Wang, O. Potapova, O. Voronina, L. Desjardins, O. Mariani, S. Roman-Roman, X. Sastre, M. H. Stern, F. Cheng, S. Signoretti, A. Berchuck, D. Bigner, E. Lipp, J. Marks, S. McCall, R. McLendon, A. Secord, A. Sharp, M. Behera, D. J. Brat, A. Chen, K. Delman, S. Force, F. Khuri, K. Magliocca, S. Maithel, J. J. Olson, T. Owonikoko, A. Pickens, S. Ramalingam, D. M. Shin, G. Sica, E. G. Van Meir, W. Eijckenboom, A. Gillis, E. Korpershoek, L. Looijenga, W. Oosterhuis, H. Stoop, K. E. van Kessel, E. C. Zwarthoff, C. Calatozzolo, L. Cuppini, S. Cuzzubbo, F. DiMeco, G. Finocchiaro, L. Mattei, A. Perin, B. Pollo, C. Chen, J. Houck, P. Lohavanichbutr, A. Hartmann, C. Stoehr, R. Stoehr, H. Taubert, S. Wach, B. Wullich, W. Kycler, D. Murawa, M. Wiznerowicz, K. Chung, W. J. Edenfield, J. Martin, E. Baudin, G. Bubley, R. Bueno, A. De Rienzo, W. G. Richards, S. Kalkanis, T. Mikkelsen, H. Noushmehr, L. Scarpace, N. Girard, M. Aymerich, E. Campo, E. Giné, A. L. Guillermo, N. Van Bang, P. T. Hanh, B. D. Phu, Y. Tang, H. Colman, K. Evason, P. R. Dottino, J. A. Martignetti, H. Gabra, H. Juhl, T. Akeredolu, S. Stepa, D. Hoon, K. Ahn, K. J. Kang, F. Beuschlein, A. Breggia, M. Birrer, D. Bell, M. Borad, A. H. Bryce, E. Castle, V. Chandan, J. Cheville, J. Copland, M. Farnell, T. Flotte, N. Giama, T. Ho, M. Kendrick, J. P. Kocher, K. Kopp, Moser, D. Nagorney, D. O’Brien, B. P. O’Neill, T. Patel, G. Petersen, F. Que, M. Rivera, L. Roberts, R. Smallridge, T. Smyrk, M. Stanton, R. H. Thompson, M. Torbenson, J. D. Yang, L. Zhang, F. Brimo, J. A. Ajani, A. M. Angulo Gonzalez, C. Behrens, J. Bondaruk, R. Broaddus, B. Czerniak, B. Esmaeli, J. Fujimoto, J. Gershenwald, C. Guo, C. Logothetis, F. Meric-Bernstam, C. Moran, L. Ramondetta, D. Rice, A. Sood, P. Tamboli, T. Thompson, P. Troncoso, A. Tsao, I. Wistuba, C. Carter, L. Haydu, P. Hersey, V. Jakrot, H. Kakavand, R. Kefford, K. Lee, G. Long, G. Mann, M. Quinn, R. Saw, R. Scolyer, K. Shannon, A. Spillane, J. Stretch, M. Synott, J. Thompson, J. Wilmott, H. Al-Ahmadie, T. A. Chan, R. Ghossein, A. Gopalan, D. A. Levine, V. Reuter, S. Singer, B. Singh, N. V. Tien, T. Broudy, C. Mirsaidi, P. Nair, P. Drwiega, J. Miller, J. Smith, H. Zaren, J. W. Park, N. P. Hung, E. Kebebew, W. M. Linehan, A. R. Metwalli, K. Pacak, P. A. Pinto, M. Schiffman, L. S. Schmidt, C. D. Vocke, R. Worrell, H. Yang, M. Moncrieff, C. Goparaju, J. Melamed, H. Pass, N. Botnariuc, I. Caraman, M. Cernat, I. Chemencedji, A. Clipca, S. Doruc, G. Gorincioi, S. Mura, M. Pirtac, I. Stancul, D. Tcaciuc, M. Albert, I. Alexopoulou, A. Arnaout, J. Bartlett, J. Engel, S. Gilbert, J. Parfitt, H. Sekhon, G. Thomas, D. M. Rassl, R. C. Rintoul, C. Bifulco, R. Tamakawa, W. Urba, N. Hayward, H. Timmers, A. Antenucci, F. Facciolo, G. Grazi, M. Marino, R. Merola, R. de Krijger, A. P. Gimenez-Roqueplo, A. Piché, S. Chevalier, G. McKercher, K. Birsoy, G. Barnett, C. Brewer, C. Farver, T. Naska, N. A. Pennell, D. Raymond, C. Schilero, K. Smolenski, F. Williams, C. Morrison, J. A. Borgia, M. J. Liptay, M. Pool, C. W. Seder, K. Junker, L. Omberg, M. Dinkin, G. Manikhas, D. Alvaro, M. C. Bragazzi, V. Cardinale, G. Carpino, E. Gaudio, D. Chesla, S. Cottingham, M. Dubina, F. Moiseenko, R. Dhanasekaran, K. F. Becker, K. P. Janssen, J. Slotta-Huspenina, M. H. Abdel-Rahman, D. Aziz, S. Bell, C. M. Cebulla, A. Davis, R. Duell, J. B. Elder, J. Hilty, B. Kumar, J. Lang, N. L. Lehman, R. Mandt, P. Nguyen, R. Pilarski, K. Rai, L. Schoenfield, K. Senecal, P. Wakely, P. Hansen, R. Lechan, J. Powers, A. Tischler, W. E. Grizzle, K. C. Sexton, A. Kastl, J. Henderson, S. Porten, J. Waldmann, M. Fassnacht, S. L. Asa, D. Schadendorf, M. Couce, M. Graefen, H. Huland, G. Sauter, T. Schlomm, R. Simon, P. Tennstedt, O. Olabode, M. Nelson, O. Bathe, P. R. Carroll, J. M. Chan, P. Disaia, P. Glenn, R. K. Kelley, C. N. Landen, J. Phillips, M. Prados, J. Simko, K. Smith-McCune, S. VandenBerg, K. Roggin, A. Fehrenbach, A. Kendler, S. Sifri, R. Steele, A. Jimeno, F. Carey, I. Forgie, M. Mannelli, M. Carney, B. Hernandez, B. Campos, C. Herold-Mende, C. Jungk, A. Unterberg, A. von Deimling, A. Bossler, J. Galbraith, L. Jacobus, M. Knudson, T. Knutson, D. Ma, M. Milhem, R. Sigmund, R. Madan, H. G. Rosenthal, A. Boussioutas, D. Beer, T. Giordano, A. M. Mes-Masson, F. Saad, T. Bocklage, L. Landrum, R. Mannel, K. Moore, K. Moxley, R. Postier, J. Walker, R. Zuna, M. Feldman, F. Valdivieso, R. Dhir, J. Luketich, E. M. Mora Pinero, M. Quintero-Aguilo, C. G. Carlotti, J. S. Dos Santos, R. Kemp, A. Sankarankuty, D. Tirapelli, J. Catto, K. Agnew, E. Swisher, J. Creaney, B. Robinson, E. M. Godwin, S. Kendall, C. Shipman, C. Bradford, T. Carey, A. Haddad, J. Moyer, L. Peterson, M. Prince, L. Rozek, G. Wolf, R. Bowman, K. M. Fong, I. Yang, R. Korst, W. K. Rathmell, J. L. Fantacone-Campbell, J. Hooke, A. J. Kovatich, C. D. Shriver, J. DiPersio, B. Drake, R. Govindan, S. Heath, T. Ley, B. Van Tine, P. Westervelt, M. A. Rubin, J. Il Lee, N. D. Aredes, and A. Mariamidze, Cancer Cell 33, 690 (2018).

31. L. De Mattos-Arruda, S. J. Sammut, E. M. Ross, R. Bashford-Rogers, E. Greenstein, H. Markus, S. Morganella, Y. Teng, Y. Maruvka, B. Pereira, O. M. Rueda, S. F. Chin, T. Contente-Cuomo, R. Mayor, A. Arias, H. R. Ali, W. Cope, D. Tiezzi, A. Dariush, T. Dias Amarante, D. Reshef, N. Ciriaco, E. Martinez-Saez, V. Peg, S. Ramon y Cajal, J. Cortes, G. Vassiliou, G. Getz, S. Nik-Zainal, M. Murtaza, N. Friedman, F. Markowetz, J. Seoane, and C. Caldas, Cell Rep. (2019).

32. R. Marcotte, A. Sayad, K. R. Brown, F. Sanchez-Garcia, J. Reimand, M. Haider, C. Virtanen, J. E. Bradner, G. D. Bader, G. B. Mills, D. Pe’Er, J. Moffat, and B. G. Neel, Cell (2016).

33. S. Li, Y. Shen, M. Wang, J. Yang, M. Lv, P. Li, Z. Chen, and J. Yang, Oncotarget (2017).

34. G. Lazaridis, V. Kotoula, E. Vrettou, I. Kostopoulos, K. Manousou, K. Papadopoulou, E. Giannoulatou, M. Bobos, M. Sotiropoulou, G. Pentheroudakis, I. Efstratiou, A. Papoudou-Bai Psyrri, C. Christodoulou, H. Gogas, A. Koutras, E. Timotheadou, D. Pectasides, F. Zagouri, and G. Fountzilas, Cancer Genomics Proteomics (2019).

35. H. M. Stern, H. Gardner, T. Burzykowski, W. Elatre, C. O’Brien, M. R. Lackner, G. A. Pestano, A. Santiago, I. Villalobos, W. Eiermann, T. Pienkowski, M. Martin, N. Robert, J. Crown, P. Nuciforo, V. Bee, J. Mackey, D. J. Slamon, and M. F. Press, Clin. Cancer Res. (2015).

36. X. Zhou, Q. Hao, and H. Lu, J Mol Cell Biol 11, 293 (2019).

37. P. Yang, C. W. Du, M. Kwan, S. X. Liang, and G. J. Zhang, Sci Rep 3, 2246 (2013).

38. S. C. Linn, H. M. Pinedo, J. van Ark-Otte, P. van der Valk, K. Hoekman, A. H. Honkoop, J. B. Vermorken, and G. Giaccone, Int J Cancer 71, 787 (1997).

39. L. S. Steelman, P. M. Navolanic, M. L. Sokolosky, J. R. Taylor, B. D. Lehmann, W. H. Chappell, S. L. Abrams, E. W. T. Wong, K. M. Stadelman, D. M. Terrian, N. R. Leslie, A. M. Martelli, F. Stivala, M. Libra, R. A. Franklin, and J. A. McCubrey, Oncogene (2008).

40. Q. Wang, Y. L. Shi, K. Zhou, L. L. Wang, Z. X. Yan, Y. L. Liu, L. L. Xu, S. W. Zhao, H. L. Chu, T. T. Shi, Q. H. Ma, and J. Bi, Cell Death Dis 9, 739 (2018).

